# Safety and Feasibility of High-density Diffuse Optical Tomography for Longitudinal Brain Monitoring in Pediatric ECMO Patients

**DOI:** 10.1101/2025.05.08.25326960

**Authors:** Sung Min Park, Sophia R. McMorrow, Tessa G. George, Chloe M. Sobolewski, Dalin Yang, Emma Speh, Evan Daniels-Day, Ari Segel, Kelsey T. King, Jeanette Kenley, Christopher D. Smyser, Joseph P. Culver, Kristin P. Guilliams, Adam T. Eggebrecht, Ahmed S. Said

## Abstract

Extracorporeal membrane oxygenation (ECMO) provides life support for severe, reversible cardiac or respiratory failure but carries substantial risk of neurological complications. Pediatric ECMO patients are particularly vulnerable, with over half of survivors exhibiting abnormal neuroimaging findings following discharge. Currently available clinical neuroimaging tools are limited, offering either static anatomical snapshots (ultrasound, computed tomography) or sparse functional monitoring (electroencephalography, functional near infrared spectroscopy) with limited spatial specificity. High-density diffuse optical tomography (HD-DOT) addresses these limitations to provide noninvasive, longitudinal, wide-field measurements of changes in cortical oxygenation at the beside. Here, we investigate safety and feasibility for bedside longitudinal HD-DOT monitoring of cerebral oxygenation focusing on data collected over 20 days in seven pediatric ECMO patients. Results confirm the reliable acquisition of high-quality HD-DOT data without adverse events, establishing HD-DOT as a promising tool for continuous, and safe bedside neuroimaging in this population.

## Introduction

Extracorporeal membrane oxygenation (ECMO) provides life-sustaining support for patients with severe, reversible cardiac and/or respiratory failure. More than 45,000 pediatric and neonatal patients have received ECMO support between the years 2009 to 2022,^1^ significantly improving survival rate in this vulnerable population. However, neurological morbidity remains high among survivors^2^, with a significant proportion of pediatric ECMO patients exhibiting functional cognitive and behavioral impairment at hospital discharge.^3,4^ One of the fundamental challenges in addressing these neurologic morbidities stems from limitations in conventional neuroimaging and neuromonitoring modalities^3^. Traditional magnetic resonance imaging (MRI) is incompatible with ECMO circuits, computed tomography (CT) involves radiation exposure, and head ultrasound (HUS) is restricted to infants with open fontanelles.^5^ Electroencephalography (EEG) is used post ECMO cannulation to detect seizures, but it has limited sensitivity to detect underlying brain injuries in the absence of seizures.^5^ As such, traditional imaging and monitoring methods are impractical to provide the continuous bedside neurological monitoring that may detect and track evolving brain injury without interruption of standard care.

High density diffuse optical tomography (HD-DOT) is an optical neuroimaging method that offers a promising alternative to overcome these limitations. The HD-DOT technology is bedside-deployable, non-invasive, and provides information on relative changes in cortical hemoglobin concentration with spatial resolution approaching that of functional MRI.^6-8^ Previous research has demonstrated initial feasibility of using bedside HD-DOT to estimate functional connectivity in neonates,^7,9^ including in a neonate undergoing veno-arterial (VA) ECMO.^10^ This study revealed stable functional connectivity during periods of clinical stability and notable disruptions during a clamp trial.^10^

In this study we aimed to evaluate the safety and feasibility of HD-DOT for longitudinal neuromonitoring in the pediatric ECMO patients supported on different modes of ECMO and with different cannulation approaches. Our secondary aim was to evaluate the association of longitudinal changes in HD-DOT acquired data representing real-time changes in cerebral oxygenation with changes with standard clinical parameters during stable and interrupted ECMO support, clamp trial.

## Materials & Methods Patient Recruitment

ECMO Patients were recruited from the St. Louis Children’s Hospital. Parental informed consent was obtained for each patient’s parent or guardian as approved by the Human Research Protections Office at Washington University School of Medicine Institutional Review Board (IRB#202205106). We recruited seven patients, including both veno-arterial (VA), and veno-venous (VV) ECMO patients. Patients underwent HD-DOT scans when deemed clinically stable by the medical team. Patients had no head wounds that would interfere with cap placement.

### HD-DOT System

We employed a continuous wave HD-DOT system optimized for bedside pediatric imaging.^10^ The system consists of 80 dual-wavelength laser diodes (685nm, 830nm) and 78 avalanche photodiode detectors (APDs), arranged to cover the bilateral parietal, occipital, temporal, and prefrontal regions (**Fig. 1A**). This configuration provides over 1200 multi-distance, overlapping measurements (≤ 4cm) with a 10 Hz sampling rate. The system components are housed in a console connected to the imaging array cap via 6 meter long, flexible fiber bundles, allowing for the console to be positioned up to 3 meters away to avoid interference with patient care. Imaging array caps, constructed from 3D-printed thermoplastic polyurethane shell lined with foam padding (**Fig. 1B**). While an individual cap may be used for a range of ∼6 cm in head circumference, caps are built to cover a range of patient head sizes from <34 cm to >60 cm. Resin-printed fiber tips defocus light and increase the optical footprint while maintaining perpendicular scalp contact. Fiber tips protrude ∼3 mm through the flexible cap to comb through hair and maintain stable optical coupling given idiosyncratic variability in scalp shape. The overall design ensures patient comfort during extended scanning sessions.

**Figure 1.**
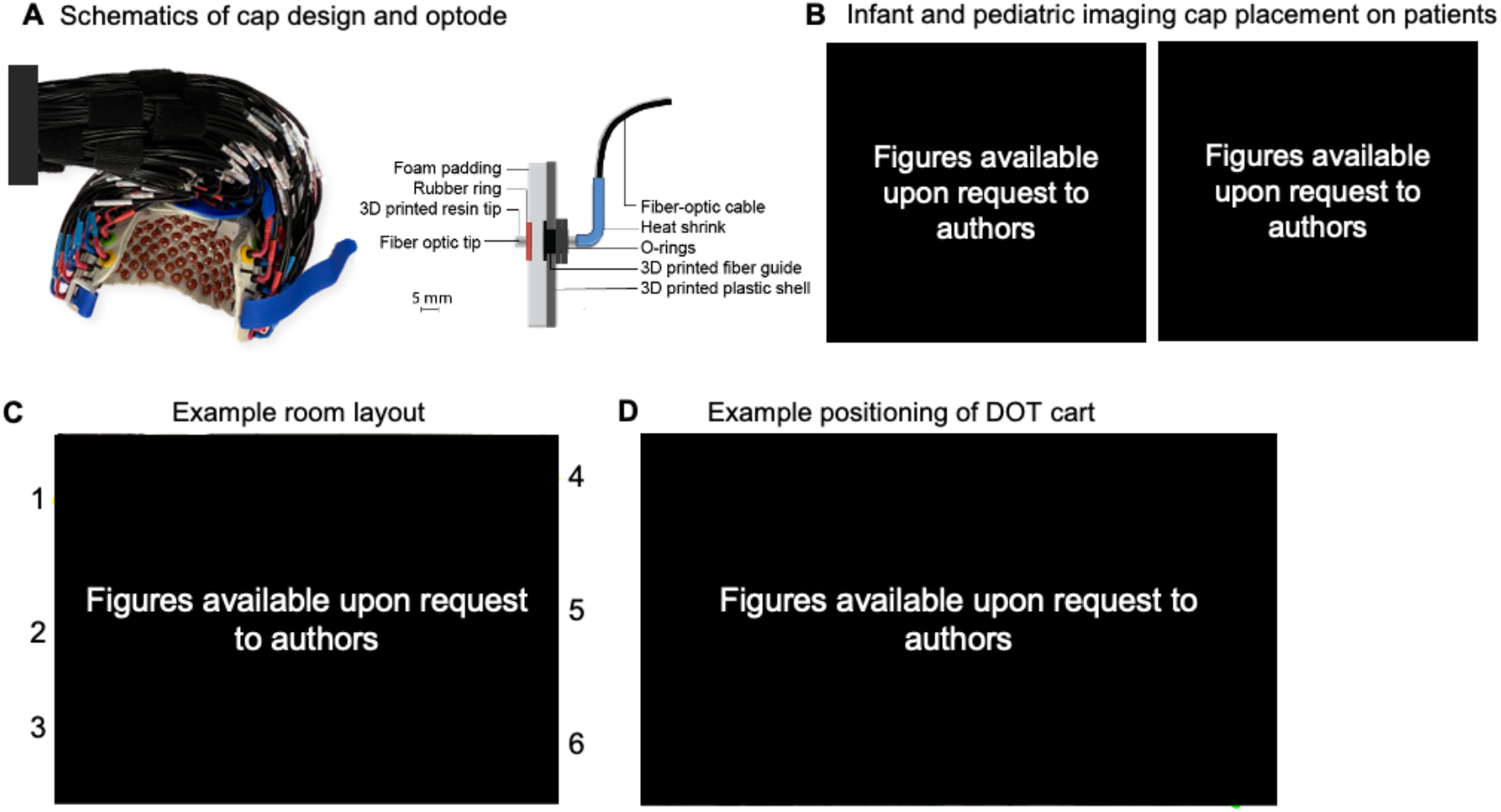
HD-DOT system components. **A** The HD-DOT imaging cap was optimized for infants (Left) and pediatric (right) populations and designed to provide comfort during extended scanning sessions. The patient’s head is positioned in the cap such that the bottom row of optodes rests just above the ears. Infant cap fits head circumferences between 34 cm to 39 centimeters, and the pediatric imaging cap was designed to fit head circumferences between 40-50 centimeters. **B.** 3D printed shell provides structural support, and foam padding and 3D printed resin tips maximize patient comfort. O-rings, 3D printed fiber guides, and rubber rings secure the optodes. **C** Example room set up with the HD-DOT system deployed in a critical care setting, including 1) vitals monitor, 2) ECMO circuit, 3) hospital bed with patient wearing HD-DOT cap, 4) supports with outlets, oxygen, medical air access, and computers, 5) HD-DOT console, 6) ventilator. **D** Long, flexible fiber optic bundles connecting the imaging cap and the cart allows for the cart, allowing the equipment to be positioned without obstructing access to patients or the ECMO circuit.

### Data acquisition

The HD-DOT cap was fitted to the patients’ heads and secured with Velcro straps without interfering with cannula position or securement. We documented cap placement with photographs taken at multiple angles for subsequent head modeling. We closely monitored the patient for any signs of discomfort. Pre- and postscan photos were taken to monitor any contusions or bruises.

Concurrent physiological measurements were obtained from standard clinical monitoring, including labs, ventilation, ECMO settings and vital signs, including heart rate (HR), mean arterial pressure (MAP), oxygen saturation (SpO2), renal near-infrared spectroscopy (NIRS), and end-tidal carbon dioxide (etCO2). Bedside measurements were recorded at 0.2 Hz, up-sampled to 1Hz and manually time-synchronized with HD-DOT data.

### Data Processing

We assessed HD-DOT data quality both during acquisition and post-collection. Real-time data quality metrics included monitoring the light levels as a function of source-detector separations (**Fig. 2A**), the pulse signal-to-noise ratio (SNR; **Fig. 2B**) and coupling coefficients for each optode in the imaging array (**Fig. 2C**). Post-collection data quality assurance analyses with NeuroDOT software focused on data with standard deviation less than 7.5% of mean signal (**Fig. 2D**) and stable optode coupling via median pulse SNR across the array (**Fig. 2E**). Noisy segments were manually excluded from further analysis.

**Figure 2.**
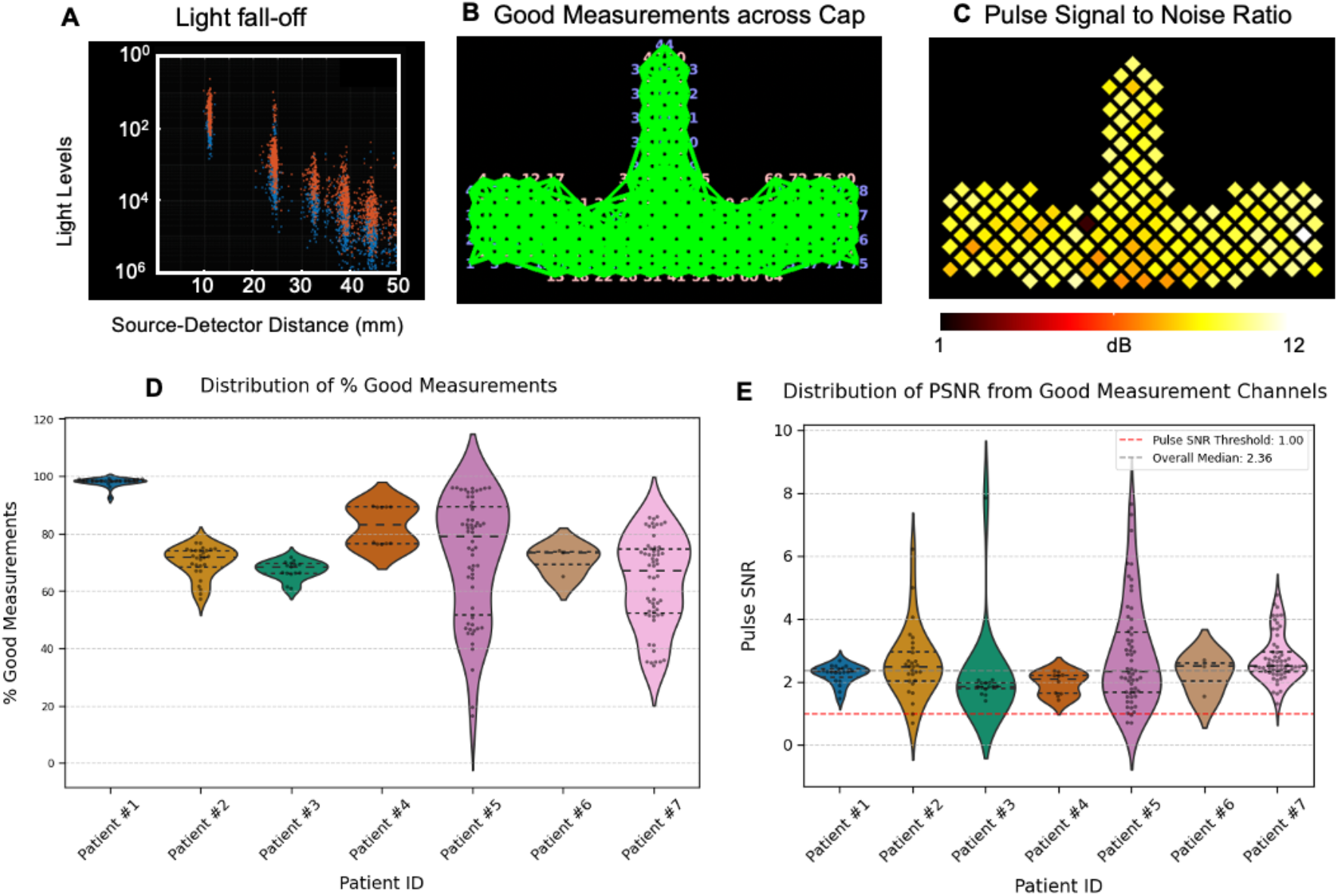
Data Quality Assessment. A Temporal mean of measurements across the cap exhibits a log-linear decay related to source-detector distances. **B** An example patient’s average coupling coefficients of each optode across the cap that remains within two orders of magnitude, indicating good data quality. **C** A high signal to noise ratio displayed on a flattened view of the cap. **D** Distribution of percentage good measurement (GM) channels across subjects, defined as percentage of channels below 35mm source-detector separation with optical density standard deviation under 7.5%. **E** Distribution of pulse signal-to-noise ratio, computed as the ratio of the integrated band limited power at the pulse frequency (within a window of 0.4Hz centered around cardiac frequency) to the median power with equivalent bandwidth in a lower frequency band.

### Cerebral and Superficial Tissue Oxygenation

Using NIRFAST software,^11^ we modeled light propagation and depth sensitivity with an age-appropriate atlas scaled to patient head dimensions (**Fig.3A-C**), with optode placement guided by facial landmarks and cap markers (**Fig. 3D**). Data were reconstructed to obtain volumetric movies of absorption (Fig. 3D).^12-14^ Spectroscopy yielded time-series of relative changes in concentration of oxyhemoglobin (ΔHbO), deoxyhemoglobin (ΔHbR), total (ΔHbT = ΔHbO + ΔHbR) and difference (ΔHbD = ΔHbO -ΔHbR) in hemoglobin species. Data were spatially smoothed (3D Gaussian kernel, 3mm full width at half maximum) and resampled to 1 Hz, spatially registered to the standard MNI space, and intersected with the Gordon parcellation^15^ to define regions of interests (**Fig. 3F-G**). Cerebral oxygenation time traces were extracted for bilateral motor and visual parcels and z-scored, while superficial tissue hemodynamics were calculated from the first nearest neighbor measurements (9 mm separation).

**Figure 3.**
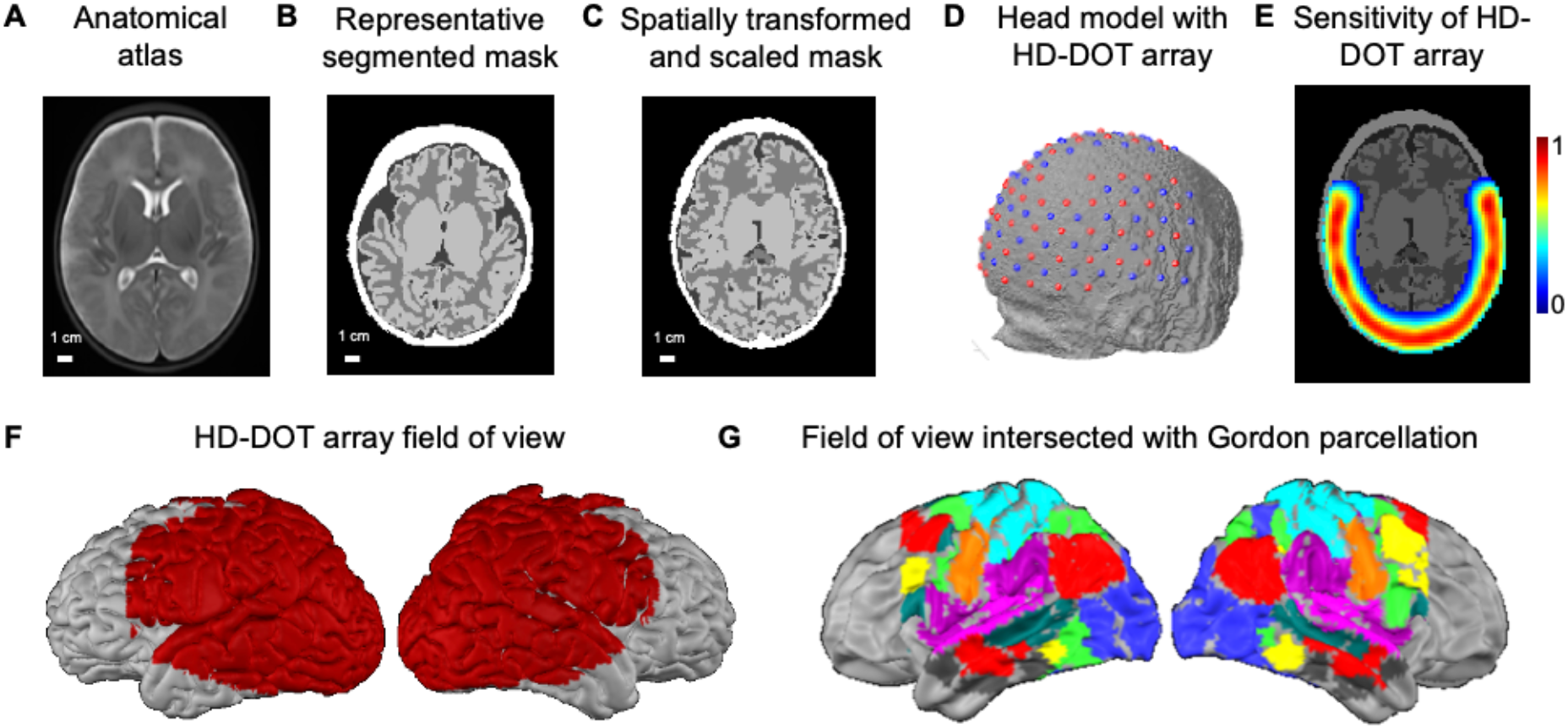
Light modeling and spatial normalization. **A** Age-appropriate anatomical atlas. **B** Representative segmented mask from an infant with comparable head circumference and age. **C** Segmented mask spatially registered to neonatal atlas space to create a head model. **D** Head mesh with HD-DOT array. **E** Depth sensitivity of the HD-DOT array field of view. **F** Resultant field of view shown in red on lateral views of the cortical surface. **G** HD-DOT array field of view intersected with the Gordon parcellation colored by putative functional networks.

## Results

### Safety and Feasibility Outcomes

The HD-DOT caps were adaptable and compatible with both central and neck cannulation sites. We successfully scanned seven neonatal and pediatric ECMO patients undergoing VA or VV ECMO (**Table 1**). We acquired data for a total of 1,915 minutes (∼32 hours) over 20 scan sessions. The average scan duration per session was 95.73 minutes (median: 99.98 minutes, range: 22.98 - 156.30 minutes). There were no incidences of scalp bruising or contusions from the fiber tip. The HD-DOT data acquisition did not interfere with ongoing clinical care procedures, including apheresis, clamp trials, sweep off trials, or respiratory therapy procedures. (**Table 2**). Importantly, no safety events associated with the HD-DOT system were reported.

**Table 1.** Patient Demographics. Demographics and Clinical Characteristics of Patients in the study. Demographic and clinical characteristics of the study population, including sex, age, head circumference (Head Circ.), ECMO type (VA: Venoarterial, VV: Venovenous), and duration of DOT data collection in minutes. ECMO = Extracorporeal membrane Oxygenation. DOT = Diffuse optical tomography.

| Patient # | Sex | Age | Head<br>Circ.<br>(cm) | ECMO Type | DOT Duration<br>(min) | Days of data<br>collection |
| --- | --- | --- | --- | --- | --- | --- |
| 1 | F | 0 to 3<br>years | 34.5 | VA | 190.7 | 2 |
| 2 | F | 12 to 16<br>years | 45.5 | VV | 268.7 | 3 |
| 3 | M | 0 to 3<br>years | 47.0 | VA | 151.4 | 1 |
| 4 | F | 0 to 3<br>years | 50.0 | VA | 97.4 | 2 |
| 5 | F | 8 to 11<br>years | 47.0 | VA | 608.7 | 5 |
| 6 | M | 0 to 3<br>years | 36.0 | VA | 23.0 | 1 |
| 7 | F | 4 to 7<br>years | 46.0 | VV | 574.7 | 6 |

**Table 2.** Procedures and Events during HD-DOT scans. Table summarizes monitoring sessions for ECMO patients, including patient ID, cumulative days on ECMO on the day of the HD-DOT scan, scan duration (minutes) and concurrent events during HD-DOT scans. Clinical events include routine examinations (inspection of chest sounds, inspection of ECMO circuit components, assessment of catheters and IV lines, pupillary examination) and any interventions or changes in patient status during HD-DOT monitoring period.

| Patient ID | Cumulative ECMO days at HD-DOT scan | HD-DOT scan duration (min) | Concurrent Events during HD-DOT scan |
| --- | --- | --- | --- |
| 1 | 3 | 130.55 | Clamp trial, sonography, neurological exam |
| 1 | 4 | 60.14 | Routine examination |
| 2 | 4 | 112.11 | Apheresis, repositioning patient, routine examination |
| 2 | 7 | 105.64 | Routine examination, dialysis, repositioning patient |
| 2 | 8 | 50.98 | Repositioning patient, diaper change, routine examination |
| 3 | 19 | 151.42 | Routine examination, physiotherapy |
| 4 | 6 | 50.50 | None documented |
| 4 | 7 | 46.97 | Respiratory therapy |
| 5 | 6 | 156.30 | Ventilator change, respiratory therapy, routine examination |
| 5 | 9 | 150.57 | Echocardiogram, routine checks, suctioning breathing tubes, repositioning patient |
| 5 | 10 | 128.71 | Diaper change, routine examination, family interaction, ventilator change |
| 5 | 11 | 101.19 | Ventilator change, routine examination |
| 5 | 12 | 71.88 | Clamp trial, routine checks, echocardiogram |
| 6 | 11 | 22.98 | None documented |
| 7 | 6 | 60.15 | Routine examination |
| 7 | 10 | 104.4 | Routine examination, diaper change, PEEP titration trial, Neurology consult |
| 7 | 13 | 98.54 | Repositioning patient, Respiratory therapy (IPV), routine examination |
| 7 | 17 | 82.83 | Sweep off trial, routine examination, suctioning breathing tubes, repositioning patient |
| 7 | 18 | 91.11 | Routine examination, repositioning patient |
| 7 | 19 | 137.68 | Sweep off trial, routine examination, respiratory therapy (IPV), neurology examination |

### Data Quality Assessment

We evaluated how well the cap was coupled to the patient’s head throughout data acquisition by calculating the good measurement (GM) percentage (for channels below 35mm source-detector separation) across all runs. Good measurement channels are given by a temporal standard deviation below 7.5%^6^. In all, 77.49% of runs had GM percentage greater than 60%, indicating robust cap-to-scalp coupling in the vast majority of the runs.

We quantified data quality using the pulse signal-to-noise ratio (SNR), calculated as the ratio of the integrated power at the pulse frequency (±0.2Hz around cardiac frequency) to the median power on either side of the cardiac frequency. The pulse SNR serves as an excellent measure of optical system-scalp coupling and directly assesses sensitivity to vascular physiology.^16,17^ Additionally, we applied a threshold of median pulse SNR values greater than 1dB based on previous pediatric and adult HD-DOT studies.^10,17^ Our analyses demonstrated that 97.35% of runs exceeded this threshold, confirming high signal quality across sessions. Among channels passing these stringent data quality thresholds, the median pulse SNR was substantially higher than the 1dB threshold (**Fig. 2E)**. Even in cases with a lower overall coupling percentage reflected in reduced GM, the subset of channels achieving good scalp contact demonstrates excellent pulse SNR with group median of 2.36dB, well above our 1dB threshold (**Fig. 2E**).

### Superficial and Cerebral Changes in Hemodynamics in Response to Clamp Trial in a Neonatal VA-ECMO Patient

To highlight the richness of the HD-DOT data, we present additional findings from the same neonatal VA-ECMO patient previously described in our previous work^10^. The patient, diagnosed with hypoplastic left heart syndrome, was supported on VA-ECMO following the Norwood procedure. On the day three of VA-ECMO support, we collected HD-DOT data before and during the clamp trial. As a proof-of-concept analysis, we explore relationships between superficial and cerebral oxygenation derived from HD-DOT and physiological parameters. During the clamp trial, SpO2, renal NIRS and MAP immediately decreased and returned towards baseline when ECMO flow resumed (**Fig. 4B**). The patient was deemed not ready to separate from ECMO and full ECMO support was resumed.

**Figure 4.**
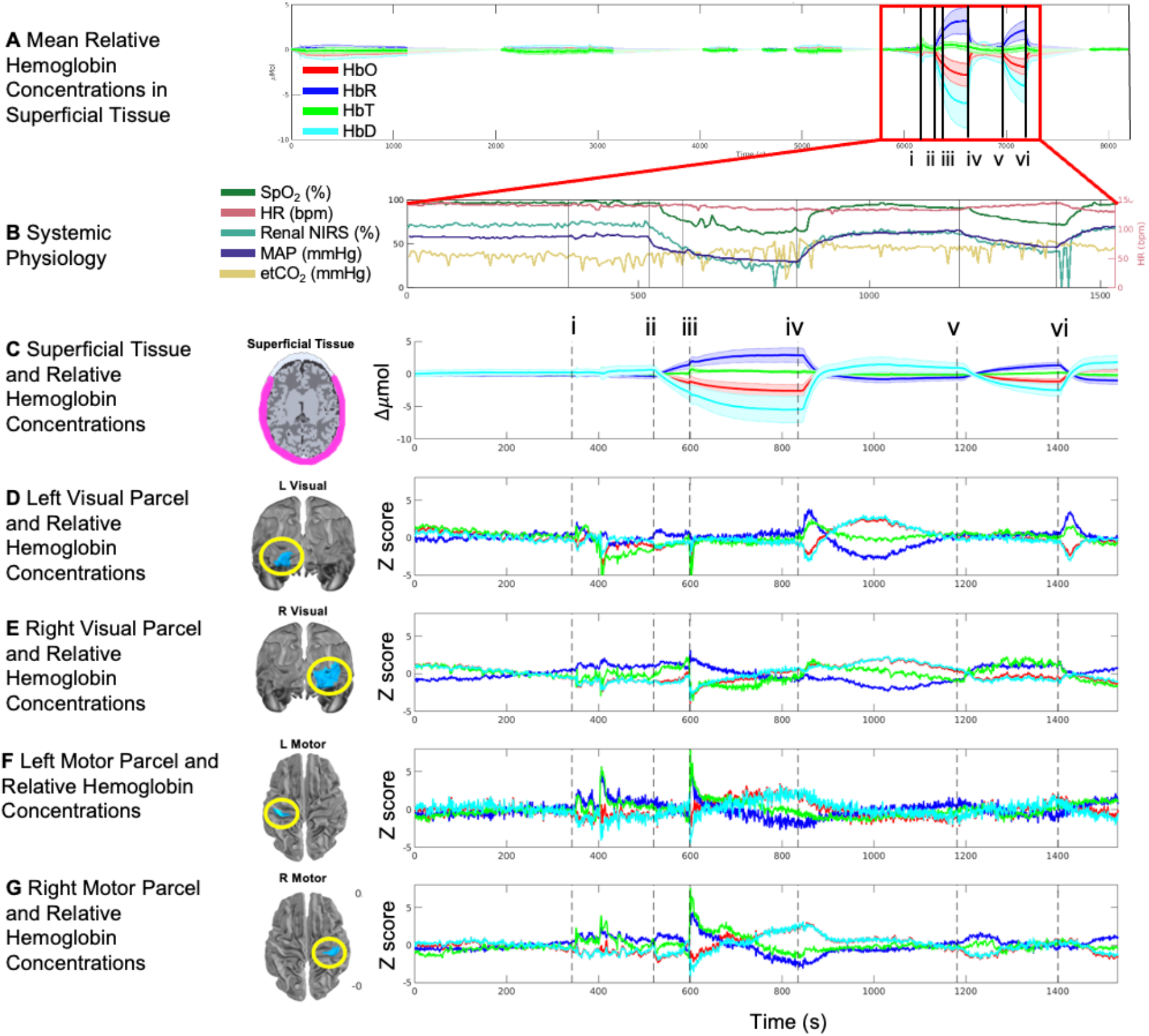
Systemic physiology, relative hemoglobin concentrations in superficial and cerebral tissue during the clamp trial. **A** Mean relative hemoglobin concentrations in superficial tissue remain consistent during baseline periods, with changes induced by clamp trial events, highlighted by red boxes (i-vi). **B** Vitals monitored as a part of standard clinical care were obtained and aligned with HD-DOT data. SpO_2_, MAP, and renal NIRS show immediate changes in response to clamp trial events. **C** Mean relative hemoglobin concentrations in superficial tissue. Shaded regions show standard deviation. Relative hemoglobin concentrations in **D** a parcel in the left visual area, **E** a parcel in the right visual area, **D** a parcel in the left motor area, and **E** a parcel in the right motor area. Events are labeled as black dotted lines, i) Mediactions administered prior to clamp trial, ii) ECMO flow stopped, iii) calcium administered, iv) ECMO resumed, v) ECMO flow stopped, vi) ECMO flow resumed.

During the clinically stable period preceding the clamp trial, superficial concentrations of HbO, HbR, HbT, and HbD remained stable. HD-DOT data revealed rapid and significant changes in relative concentrations of HbO, HbR, and their difference, HbD, during the clamp trial (**Fig. 4A**). When ECMO flow was paused, HbR rapidly increased while HbO and HbD decreased, with all metrics returning to baseline levels upon ECMO flow resumption. A similar pattern was observed during subsequent interruptions of ECMO flow, with HbR increasing immediately and HbO and HbD decreasing. HbT remained relatively stable in superficial tissue, compared to other hemoglobin concentrations. Additionally, at approximately 600 seconds, a small perturbation in hemoglobin concentrations (HbO, HbR, HbT, HbD) coincided with intravenous administration of calcium.

Cerebral hemoglobin concentrations exhibited responses to the clamp trial that were distinct from that observed in superficial tissue (**Fig. 4C-G**). Time traces of hemoglobin concentrations related to left and right parcels in visual and motor regions exhibited variable responses to the clamp trial events. The left motor and left visual parcels showed perturbations in hemoglobin concentrations at 400 seconds and 600 seconds, coinciding with medication administration before the clamp trial. The variability between superficial and cerebral tissues, as well as regional variability within the cerebral tissue highlights the spatial specificity of cerebral hemodynamics captured by wide-field HD-DOT during acute changes in physiology due to clamp trial.

## Discussion

Neuromonitoring in pediatric ECMO patients remains challenging due to limitations in clinically available neuroimaging tools. In this study, we demonstrate the safety and feasibility of HD-DOT via data quality results from seven pediatric and neonatal ECMO patients over several data collection session per patient and for extended durations. Our safety outcomes confirm successful longitudinal data collection across a diverse patient demographic without adverse events during over 1,915 minutes of data acquisition. We effectively acquired measurements from patients with different cannulation configurations (central or peripheral) without compromising clinical care. In one pediatric VA ECMO patient with peripheral cannula positioned behind the ears, we adapted the HD-DOT cap by strategically removing a subset of optodes, allowing for safe cannula placement while maintaining good data quality (Fig. 1B).

Optical imaging offers a minimal-risk, portable, and continuous alternative for functional neuromonitoring, with recent studies highlighting its potential utility for bedside monitoring in ECMO patients.^10^ While not directly measuring cerebral blood flow (CBF), multiple studies have used oxygenated hemoglobin concentration measured by near infrared spectroscopy (NIRS) to assess cerebral autoregulation (CA).^18,19^ Research using multichannel NIRS on pediatric ECMO patients, examined correlation between cerebral oxyhemoglobin (HbO) and MAP to assess CA, with notable regional variations between left and right hemispheres.^19^ Another study demonstrated that patients with an acute neurological event had a higher cerebral oxygenation index and remained outside of optimal ranges of the cerebral oxygenation index for longer periods of time during ECMO support.^18^

Other optical neuroimaging methods, including diffuse correlation spectroscopy (DCS) and frequency domain diffuse optical spectroscopy (FD-DOS), have been employed to measure cerebral blood flow (CBF) and cerebral oxygen saturation, respectively^20^. This study reported that MAP and pump flow correlated weakly with CBF and was not correlated with cerebral oxygen saturation, suggesting that systemic measures alone may be insufficient surrogates for brain health in clinical decision-making. Although research on optical neuromonitoring methods for ECMO patients is still growing, these techniques may provide invaluable insights into brain health that can further inform clinical decisions.

While the optical methods discussed above show great promise in assessing local dynamics of cerebral oxygenation or blood flow, they are limited by small fields of view. The lack of spatial coverage and spatial resolution may result in missed neurological injury that could be detected with wide-field imaging methods. Recent developments in HD-DOT allow for mapping spatially distributed brain function with image quality approaching that of functional MRI^6,13,21-23^. HD-DOT offers improved image quality and reliability over traditional fNIRS^6^. Our recently published case study assessing functional connectivity metrics^10^ and the results on the spatial variability of cerebral oxygenation during acute physiological changes presented here further emphasize the need for functional neuromonitoring tools that interrogate a wide field of view.

In our recently published work, we demonstrated the feasibility of assessing functional connectivity at the individual patient level, revealing global disruption of low bandwidth correlation between brain regions during clamp trials. This study further demonstrates spatial and temporal variability in cerebral oxygenation and its relationship with systemic physiology.

Relative hemoglobin cerebral concentrations in superficial and cortical tissue showed minimal variability during baseline but exhibited pronounced disruptions during clamp trials. Notably, responses in HbO and HbD from superficial tissue differed from that of cortical tissue but resembled the simultaneously recorded changes in SpO_2_, MAP, and renal NIRS during clamp trial. This differential response pattern may be attributed to cerebral autoregulation. Spatially variable cortical oxygenation patterns were also observed during the clamp trial (Fig. 4C-G), with distinct changes in different brain regions. While further research is necessary to understand the underlying physiology driving these regional variations, our HD-DOT system’s extended field of view provides substantial advantages over single and sparse optode arrays.

Previous NIRS studies have examined hemoglobin concentration changes and their relationship with physiological measures in ECMO patients during pump flow manipulations.^19,24,25^ These studies documented not only differences between cerebral and peripheral tissue oxygenation^25^, but also regional differences between the left and right hemispheres.^19^ Our findings align with these observations; the ECMO patient in our study exhibited distinct cerebral oxygenation responses to stopping of ECMO flow compared to superficial tissue. Consequently, conventional systemic measures may be insufficient for guiding neuroprotective clinical decisions. HD-DOT has the potential to address these gaps by providing wide-field, spatially resolved measures of brain health that complements existing monitoring approaches.

In addition to the response to the clamp trial and changes in ECMO flow, cerebral oxygenation also showed distinct responses to medications administered during the ECMO course (**Fig. 4C**). A variety of medications were administered prior to and during the clamp trial, including sedatives. Cerebral oxygenation showed a distinct response coinciding with the administration of these medications. ECMO patients are a complex population and are often given a variety of medications that have the potential to alter cerebral blood flow, including but not limited to sedatives, analgesics, and anticoagulants. Our data show a potential response in cerebral oxygenation in response to medications. Although it is difficult to draw conclusions based on one neonatal ECMO patient, these results highlight the richness of these data and the potential of HD-DOT for monitoring a variety of clinically relevant information during EMCO support.

The use of atlas-based head modeling, while validated in previous studies,^7,26,27^ are known to have higher localization errors compared to subject-specific approaches. The optode locations were approximated using photographs, cap markers, and facial landmarks, but this can potentially introduce deviation affect downstream analyses post reconstruction. Incorporating patient-specific head anatomy via emerging photogrammetric approaches will improve spatial accuracy and measurement reliability.

A limitation of this study is the small sample size (N=7), which significantly constrains the generalizability of our findings. However, these 7 patients cover a wide range of ages, sizes, and skin tones. ECMO patients represent a complex, heterogenous population with varying underlying pathophysiology, treatment protocols, and comorbidities. Our preliminary results, while promising, need to be interpreted cautiously as they may not fully capture the variability of cerebral hemodynamic responses to across different patient population. However, the large total data collection time over multiple patients highlight both the safety and feasibility for applying HD-DOT at the clinical bedside in acute care settings.

Additionally, our study did not include long-term follow-up assessments or correlation with neurodevelopmental outcomes, limiting our understanding of the clinical significance of the observed patterns of cerebral oxygenation. Future studies will investigate the relationships of HD-DOT derived metrics with neurological assessments and patient outcomes to examine whether HD-DOT metrics can serve as predictive measures of brain health and neurological prognosis.

Our ongoing study and HD-DOT data collection efforts aim to address the sample size limitation. This larger cohort will enable more robust statistical analysis and allow for investigation on the effect of key ECMO variables on cerebral oxygenation. Longitudinal follow-up including neurological assessments and neuroimaging for patients will be essential to establish the relationship between HD-DOT measurements and both short-term outcomes at discharge and long-term neurodevelopmental trajectories. This quantification could potentially identify early markers of neurological injury. Finally, recent technological advancements in HD-DOT hardware^28^ may facilitate the development of increasingly portable and wearable systems that could expand clinical applications of HD-DOT beyond ECMO to other critical care settings requiring neuromonitoring.

## Conclusion

This study demonstrates the safety and feasibility of using HD-DOT for continuous, non-invasive, bedside neuroimaging in a pilot pediatric ECMO cohort. Through successful, longitudinal data collection over 20 days across seven patients without adverse events, we establish HD-DOT as a promising tool for assessing cerebral oxygenation. Our findings reveal spatially distributed patterns of cerebral oxygenation during ECMO that traditional neuroimaging modalities cannot capture. This work addresses a gap in neuromonitoring for pediatric ECMO patients and establishes a foundation for further understanding of cerebral physiology during extracorporeal support, and ultimately contribute to improved neurological outcomes in this high-risk population.

## Data Availability

All data produced in the present study are available upon reasonable request to the authors

## Acknowledgements

We would like to express our gratitude to the patients and families who participated in the study. All study procedures were approved by the Human Protections Office at Washington University School of Medicine Institutional Review Board (protocol #202205106). Written informed consent was obtained by each patient’s parents or legal guardians. This study was supported by funding from the Children’s Discovery Institute at St. Louis Children’s Hospital (CDI-II-2021-953-2) and the American Heart Association (AHA 25PRE1194156, AHA 25CSA1421325).

